# Childhood obesity prediction from nationwide health records

**DOI:** 10.1101/2020.11.09.20228247

**Authors:** Hagai Rossman, Smadar Shilo, Shiri Barbash-Hazan, Nitzan Shalom Artzi, Eran Hadar, Ran D. Balicer, Becca Feldman, Arnon Wiznitzer, Eran Segal

## Abstract

Rapid rise in childhood obesity prevalence worldwide and its major implications for health warrant the development of better prevention strategies. Here, we analyzed electronic health records of children from Israel’s largest healthcare provider from 2002 to 2018 and developed a model for predicting children’s obesity. Data included demographics, anthropometric measurements, medications, diagnoses, and laboratory tests of children and their families. Obesity was defined as body mass index (BMI) ≥95th percentile for age and sex. To identify the most critical time-window in which the largest annual increases in BMI percentile occurs during early childhood, we first analyzed childhood BMI trajectories among 382,132 adolescents. Next, we devised a prediction model targeted to identify children at high risk of obesity prior to this time-window, predicting obesity at 5-6 years of age based on data from the first 2 years of life of 136,196 children. Retrospective BMI analysis revealed that among obese adolescents, the greatest acceleration in BMI percentiles occured between 2-4 years of age. Our model, validated temporally and geographically, accurately predicted obesity at 5-6 years old (auROC of 0.804). Discrimination results on subpopulations demonstrated its robustness across the pediatric population. The model’s most influential features included anthropometric measurements of the child and family. Other impactful features included ancestry and pregnancy glucose. Antibiotics exposure in utero and during the first 2 years of life had no observed impact on obesity prediction. Our model, targeted to identify children prior to the most critical time-window of BMI acceleration, may allow a more accurate identification and implementation of early prevention strategies for children at high risk of obesity and can be readily embedded into healthcare systems.

## Introduction

In the past decades, the prevalence of childhood obesity has rapidly increased worldwide. A global analysis demonstrated that in 2016, 50 million girls and 74 million boys worldwide were obese^1^, making it a global public health crisis^2,3^. Predicting excess weight gain in children is important for numerous reasons. First, pediatric obesity is a multisystem disease that can greatly impact a child’s physical and mental health^4^. It is associated with a greater risk for premature mortality^5^ and earlier onset of chronic disorders such as hypertension, dyslipidemia, ischemic heart disease and type 2 diabetes, with insulin resistance identified in obese children as young as 5 years of age^6–8^. Second, previous studies demonstrated an underestimation of obesity by both parents and physicians^9,10,11^ and there is currently little guidance for healthcare professionals to identify infants at risk^12^. Finally, there is evidence supporting young age as a critical time period for intervention, which includes promoting healthy eating habits, physical activity and reducing sedentary behaviors, which are associated with more beneficial long-term outcomes^13^. While currently not many studies analyzed the effectiveness of family level interventions designed to prevent obesity in children younger than 5 years of age, current evidence suggests that behaviors which contribute to obesity can be positively impacted at an early age^14^. All of the above contributes to the value of a prediction model that can identify children at high risk for obesity at an early age. Although clinical prediction models are becoming increasingly prevalent in medicine^15^, very few studies have focused on childhood overweight or obesity prediction^16,17^ which is emerging as a global health epidemic. The current model was based on a population of children from birth to 2 years of age, and predicted their future obesity risk at 5-6 years of age using personal and familial data from their Electronic Health Records (EHR).

## Methods

### Data

We extracted features and outcomes from a nationwide dataset encompassing over 10-years of full administrative and clinical data from EHRs of an integrated health care system in Israel, Clalit Health Services. Clalit serves as a non-profit integrated care organization comprising over four million patients in Israel, more than half of the country’s population, and has a membership turnover of less than 2% annually^18^. The dataset included demographic data, weight and height measurements, clinic and hospital diagnoses, medication dispensed, and laboratory tests from 2002 to 2018 (Section 1 in the Supplementary Appendix).

### Study design and population

#### Acceleration of BMI in early childhood

First, to assess the dynamics of BMI changes in early childhood, we analyzed historical data from children who had a BMI measurement at 13-14 years of age, which is a routine weight checkup in Israel, and had at least two BMI measurements within a 1-year interval up to 13 years of age. There were 382,132 children with 1,401,803 measurements who met both criteria.

#### Childhood obesity prediction model

Second, a cohort was extracted for the prediction model that contained 882,987 children who were born between 2003 to 2013. Of them, 174,250 children had weight and length measurements during at least two routine infant checkups, which are scheduled for all Israeli infants at ages 1, 2, 4, 6, 9, 12, and 18 months. The children were further divided according to date of birth: 143,040 children were born between 2003-2012 and 31,210 were born between 2012-2013. Of them, only children who had a BMI outcome measurement between the ages of 5 to 6 years old, which is also performed routinely in Israel, were included in the cohort. Overall, 108,416 children who were born between 2003-2012 and 27,780 born between 2012-2013 were included in the cohort as training and temporal validation set respectively (Fig S1). Extracted features included maternal, paternal and siblings’ data. Overall, 133,811 children included in the cohort had maternal data, 110,280 had paternal data and 70,735 had available EHR data of at least one sibling. The characteristics of the study cohort are presented in Table 1.

**Table 1.** Characteristics of the Study Cohort.

|  |  | **Train set**<br>**(n=108,416)** | **Temporal**<br>**validation set**<br>**(n=27,780)** | **All (n = 136,196)** |
| --- | --- | --- | --- | --- |
| Obesity status at last infant routine checkup (prior to 2 years of age) | Underweight (%) | 10,045 (9.2%) | 2,327 (8.4%) | 12,327 (9.1%) |
|  | Normal weight (%) | 79,701 (73.5%) | 20,742(74.7%) | 100,443 (74%) |
|  | Overweight (%) | 11,177 (10.3%) | 2,835(10.2%) | 14,013 (10.3%) |
|  | Obese (%) | 7,493 (6.9%) | 1,875 (6.8%) | 9,368 (6.9%) |
| Number of available infant routine checkup measurements | Mean (std) | 5.1 (1.7) | 4.5 (1.8) | 4.6 (1.8) |
| Sex | Female (%) | 52,733 (49%) | 13,458 (48%) | 66,191 (49%) |
|  | Male (%) | 55,683 (51%) | 14,322 (52%) | 70,005 (51%) |
| Number of children with **maternal data** | Count (%) | 106,541 (98%) | 27,270 (98%) | 133,811 (98%) |
| Maternal age at childbirth [years] | Mean (std) | 30.1 (5.2) | 30.5 (5.2) | 30.1 (5.2) |
| Maternal pre-pregnancy BMI [m/kg<sup>2</sup>] | Mean (std) | 23.6 (4.7) | 23.3 (4.4) | 23.5 (4.6) |
| Number of children with **paternal data** | Count (%) | 88,084 (81%) | 22,196 (80%) | 110,280 (81%) |
| Paternal age [years] | Mean (std) | 33.1 (5.9) | 33.3 (5.7) | 33.2 (5.9) |
| Paternal BMI [m/kg<sup>2</sup>] | Mean (std) | 25.9 (4.4) | 25.6 (4.2) | 25.9 (4.3) |
| Number of children with **sibling data** | Count (%) | 55,070 (51%) | 15,665 (56%) | 70,735 (52%) |
| Number of siblings per child | Mean (std) | 1.1 (1.3) | 1.3 (1.4) | 1.2 (1.3) |
| Sibling BMI CDC z-score | Mean (std) | 0.0 (1.1) | -0.1 (1.1) | 0.0 (1.1) |

Abbreviations: BMI - body mass index, CDC- Center for Disease Control and Prevention, std - standard deviation

### Features

All EHR data available were binned into time periods and statistical measures (e.g., median, maximum, slope between two time points) were taken as features for each period. Dispensed medications and clinical diagnoses were categorized by ATC codes^19^ and ICD9 diagnosis codes, respectively, and counts in different time periods were taken as features. Weight, height, Weight-for-Length (WFL) and BMI data were converted to reference z-scores provided by the Center for Disease Control and Prevention (CDC)^20^. Valid measurements were defined as being in the range of 5 CDC standard deviation scores for weight and height. Features from maternal pregnancy were binned in alignment with the routine pregnancy tests schedule in Israel. Specific features of interest such as ancestry, and socioeconomic status surrogates were formulated manually. Feature generation methods are described in Section 3 in the Supplementary Appendix. Altogether, 1,556 features were devised for each child.

### Outcome

The outcome for our models was the obesity status of children at 5 to 6 years of age. We defined obesity status in accordance with health care professionals in Israel, using the CDC BMI reference percentiles. Cutoffs for normal weight, overweight, and obesity were determined using the CDC’s standard thresholds of the 85th percentile for overweight and 95th percentile for obesity. A previous study showed that using other percentile curves such as The World Health Organization (WHO) WFL, and WHO BMI provided similar estimates of obesity risk as the CDC percentiles at 5 years of age^21^. Distribution of weight status at the target age is described in Section 4 in the Supplementary Appendix, Table S1.

### Statistical analysis

#### Childhood obesity prediction model

For our prediction task, we trained Gradient Boosting trees^22^. Trees allow nonlinear and multiple feature interactions to be captured, which may be important in obtaining an accurate prediction model. The parameters of the model were tuned using cross-validation on the training set. The chosen parameters and handling of missing values are described in Section 5 in the Supplementary Appendix. As stringent tests, we used both temporal and geographical validations, thus testing the performance of our model for distribution shifts over time and geographic location. The temporal validation set contained children born between 2012-2013. The geographical validation set contained all the clinics in the most populated and multiethnic city in Israel, Jerusalem. Unless stated otherwise, the reported results are on the temporal validation sets. Full results are available in Table 2.

**Table 2.** Prediction Results.

| Table 2. Prediction Results |  |  |  |  |  |  |  |
| --- | --- | --- | --- | --- | --- | --- | --- |
| Descrimination on validation sets |  |  |  |  |  |  |  |
|  | Temporal validation set |  |  | Geographical validation set |  |  |  |
|  | auPR | auROC |  | auPR | auROC |  |  |
| Baseline | 0.226 (0.196-0.273) | 0.749 (0.718-0.779) |  | 0.154 (-0.006-0.460) | 0.725 (0.490-0.949) |  |  |
| Model | 0.307 (0.280-0.341) | 0.804 (0.779-0.828) |  | 0.217 (0.029-0.509) | 0.785 (0.601-0.960) |  |  |
| Effects of varying decision threshold on model performance |  |  |  |  |  |  |  |
|  | Predicted probability threshold |  |  |  |  |  | Baseline |
|  | 2% | 5% | 10% | 20% | 30% | 40% |  |
| Sensitivity | 0.959 | 0.799 | 0.59 | 0.367 | 0.24 | 0.148 | 0.281 |
| Specificity | 0.273 | 0.65 | 0.837 | 0.945 | 0.976 | 0.991 | 0.949 |
| PPV | 0.09 | 0.146 | 0.214 | 0.334 | 0.428 | 0.539 | 0.291 |
| NPV | 0.989 | 0.977 | 0.965 | 0.952 | 0.945 | 0.939 | 0.946 |
| Net Benefit | 0.053 | 0.039 | 0.024 | 0.013 | 0.007 | 0.004 |  |

Abbreviations: auPR/auROC - Area under the PR/ROC curve, PR - Precision-Recall, ROC -Receiver-Operator-Characteristic, NPV- Negative predictive value, PPV- positive predictive value

In order to compare the computational model to current practice, we considered the last WFL percentile routine checkup measurement available before 2 years of age as a baseline model. This is in line with current guidelines that recommend clinicians to assess a child’s current nutritional and obesity status by calculating WFL percentile or BMI percentile in children 0 to 2 years of age, or older than 2 years of age, respectively^23^. The WFL percentile thus emulates the information a caregiver has today to assess the current obesity status and future obesity risk of children younger than 2 years of age^24^. This variable also contains information of sex and age, as it standardizes by them.

An important aspect of evaluating risk probability models is clinical usefulness. The model outputs calibrated continuous risk probabilities, and applying a clinical decision thereafter (for example, a nutritional intervention) will vary and depend on the costs and benefits of the action, often assessed both clinically and economically. Decision curves^25^ offers a graphical tool to analyze clinical utility of adopting a new risk prediction model. The curves contain information that can guide clinicians to make decisions based on the risk thresholds and tradeoffs (costs and benefits) of their decision to treat. The costs and benefits can be translated into a function of the optimal threshold probability (Section 9 in the Supplementary Appendix).

We next analyzed the discrimination results measured by the area under the Precision-Recall curves (auPR)^26^ of our model across different subgroups of the study cohort. These included sex; obesity status at baseline as defined by the last available WFL percentile prior to 2 years of age; and the presence of at least one complex chronic disease, defined as at least one diagnosis from a previously defined classification system^27^.

#### Risk factor analysis from the prediction model

Finally, we investigated risk factors from the model by analyzing which features most attribute to the model’s prediction. To this end, we used the recently introduced SHAP (SHapley Additive exPlanation) method^28–30^ that aims to interpret the output of a machine learning model. A feature’s Shapley value represents the average change in the model’s output by conditioning on that feature when introducing features one at a time over all feature orderings. Shapley values are calculated individually for every child’s feature. An important property of Shapley values is that they are additive, meaning that the Shapley values of a child’s features add up to the predicted log-odds of obesity for that child. We transformed this value for each feature and child and obtained a relative risk score. We can therefore analyze feature attributions at the individual level, by examining plots of the Shapley value as a function of the feature value for all individuals (Section 8 in the Supplementary Appendix). This method enables us to capture non-linear and continuous relations between a feature’s impact on the prediction and the feature’s value. Of note, a vertical spread in such a plot implies interaction with other features in the model, which would not have been attainable using a linear model, without explicitly modelling the interaction.

Building a model with many correlated features (e.g. a child’s weight measurement at adjacent timepoints) is bound to suffer from severe collinearity of the features, and consequently the feature attributions will be spread across these related features in a non-trivial way. To tackle this, we made use of the additive property of Shapley values. By adding up the Shapely values of related features, grouped together according to domain knowledge, we could report an analysis on this group of features (Section 8 in the Supplementary Appendix). This gives better estimates of relevant risk scores. Another use of the additive property allows adding features according to groups and analyzes the model globally by taking the mean over absolute Shapely values of all children in each group of features. This gives insight on the impact of a feature group as a whole.

## Results

### BMI trajectories in early childhood

Retrospective BMI trajectories in early childhood were initially analyzed to construct a model targeting the most critical time window in which maximal acceleration of weight occurs. We divided 382,132 adolescents into two groups:obese and non-obese at 13-14 years of age **(Fig. 1A, B)**. Among those who were non-obese, the mean change in BMI z-score of children who were not obese between 1 to 14 years of age remained close to 0, with an annual mean change of less than 0.1 z-scores. However, for obese children at 13-14 years of age, the BMI z-score incremented throughout infancy and early childhood with the largest annual increase in BMI percentile observed at 2-4 years of age. This result motivated us to construct a model aimed at identifying children prior to the age of 2 years-old who will be at high risk for obesity within the subsequent 3-4 years of age.

**Figure 1.**
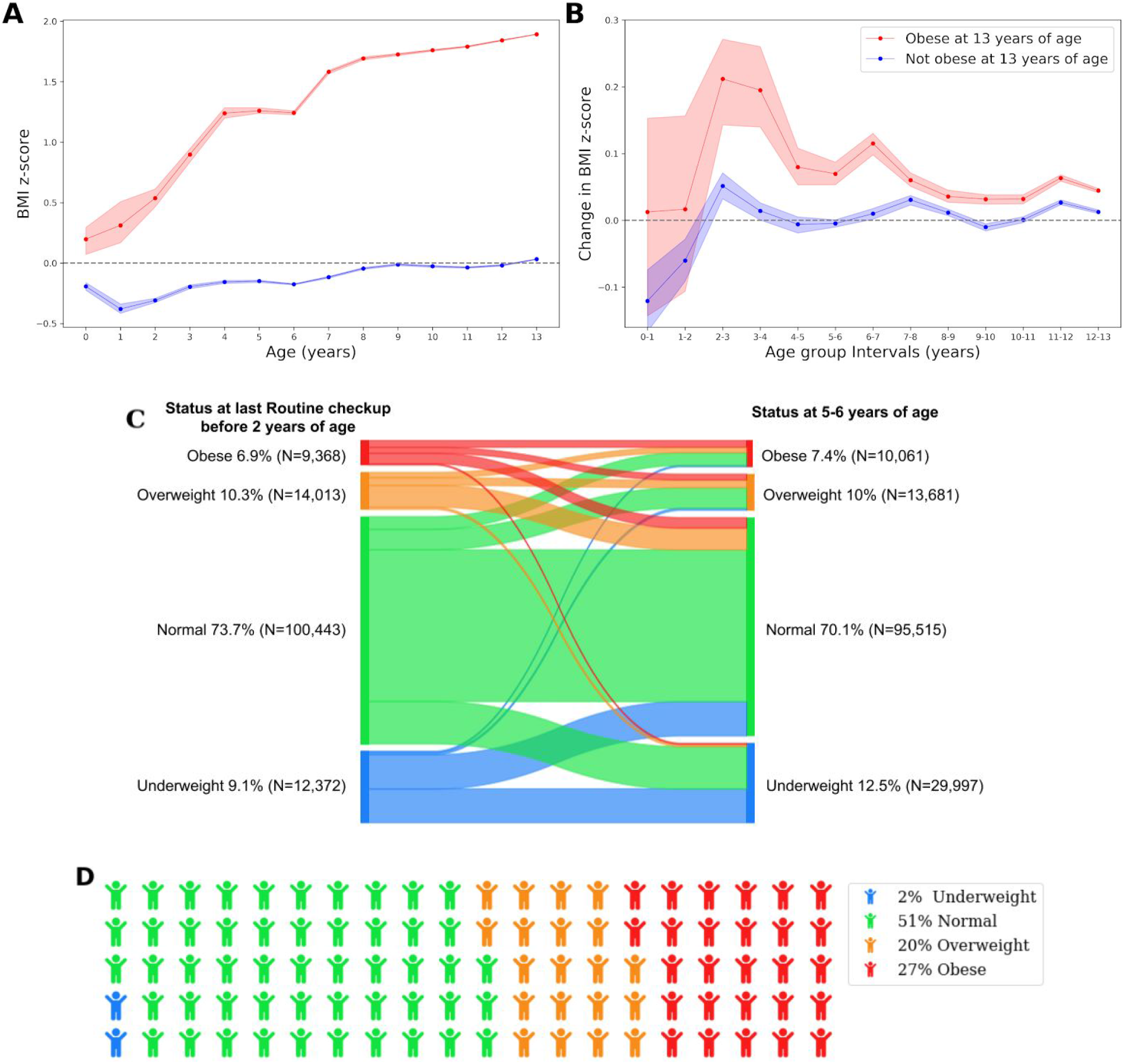
BMI dynamics in early childhood. **A:** Mean WFL/ BMI z-score for children 0-2 and 2-18 years of age respectively, who were obese (red) versus non obese (blue) at 13 years of age. **B:** Mean change in annual BMI-scores for the same groups of children. Shaded areas are 95% bootstrapped confidence intervals. **C:** Obesity status transition of the study cohort. Left side: distribution of obesity status at the last available routine checkup before 2 years of age. Right side: distribution of obesity status at 5-6 years of age. Transitions from different obesity states between these two time points are presented; Obese (red), Overweight (orange), Normal weight (green), Underweight (blue). **D:** Distribution of obesity status at infancy for obese 5-6 years old children. Abbreviations: BMI - body mass index, WFL- weight for length

Next, to further understand the BMI transition over the first 6 years of life, we analyzed the transition of obesity status in the 136,196 children that were included in our cohort. Obesity status was defined for each child at two time-points: the last available routine checkup before 2 years of age and at 5-6 years of age **(Fig. 1C)**. This analysis revealed that over half of obese children at 5-6 years of age had normal weight at infancy (51%) **(Fig. 1D)**.

### Prediction of Childhood Obesity at 5-6 years of age

We constructed a model among 136,196 eligible children age 0 to 2 years of age for prediction of childhood obesity at 5 to 6 years of age (see Methods) and evaluated the discrimination performance using the areas under the receiver operating characteristic and precision-recall curves (auROC, auPR) **(Fig. 2 A,C)**. Notably, our model outperforms the baseline model based on the child’s last WFL percentile with an auPR of 0.307 (0.280-0.341) compared to 0.223 (0.209-0.235). Both temporal and geographical validation results are summarized in Table 2. We analyzed clinical utility by constructing decision curves **(Fig 2D)**. Notably, our model dominates over other strategies in net benefit over all threshold probabilities, with significant margins in the lower threshold probability regime. A summary of the effect of applying different decision thresholds on the model performance is presented in Table 2.

**Figure 2:**
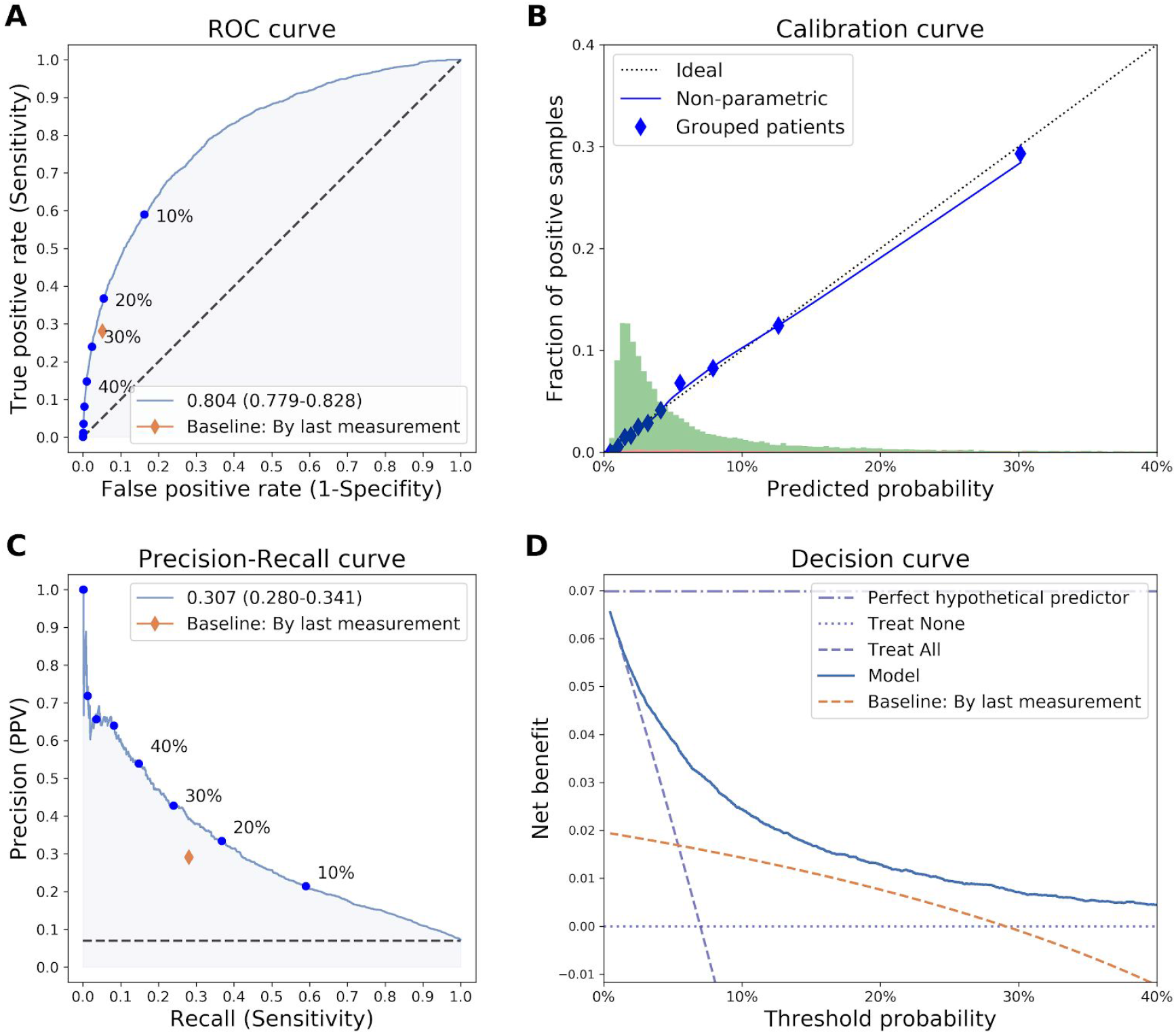
Evaluation of obesity prediction model. **A:** ROC curve of our model (blue) and a baseline model based on the last available WFL routine checkup measurement (red diamond). The blue dots and percentages represent different decision probability thresholds. **B:** Calibration curve. Blue dots represent deciles of predicted probabilities. Dotted diagonal line represents an ideal calibration. Histogram at the bottom: predicted probabilities of normal-weight children (green) and obese children (red). **C:** Precision-Recall curve. Baseline model is marked with a red diamond. Threshold percentiles are marked on the curves. **D:** Decision curve analysis containing different treatment strategies of our model (blue) and the Baseline model (red). Strategies of treating all, treating none and the perfect hypothetical predictor are also presented. Abbreviations: auPR/auROC - Area under the PR/ROC curve, PPV - positive predictive value, PR - Precision-Recall, ROC - Receiver-Operator-Characteristic, WFL - Weight-for-Length

We further analyzed the discrimination results (auPR) of our model on different subpopulations of children (section 6 in the Supplementary Appendix, Fig S6, S7). First, we evaluated the effect of sex on the model’s performance and found similar results for boys and girls. We then evaluated children who had at least one diagnosis of a complex chronic condition using a previously defined classification system^27^. The discrimination of the model was similar for children with chronic medical conditions, demonstrating the robustness of the model across a clinically heterogeneous pediatric population. Finally, discrimination performance was evaluated in subpopulation defined by obesity status as defined by the last available WFL percentile prior to 2 years of age. Our model had the highest auPR in children who were obese at infancy, followed by overweight and normal weight at infancy. It outperformed the baseline model in predicting future obesity in all infants, regardless of obesity status at baseline. Expectedly, an increase in the number of documented anthropometric measurements during routine checkups improved the discrimination performance of the model.

We next assessed the performance of multiple prediction models for childhood obesity at 5-6 years of age at the following timepoints: birth (up to the time of delivery, without birth weight and mode of delivery), 6 months, 1 year and 1.5 years of age. The effect of the child’s age at prediction and the model discrimination performance are presented in Fig. 3A and Table S2 (Section 7 in the Supplementary Appendix). As expected, the model performance improved when the prediction is done at an older age, which is closer to the age at the defined outcome. However, a prediction model based on features available up to day of birth has an auROC of 0.715 and auPR of 0.178, slightly better than the baseline model based on the child’s WFL at 1 year of age which has an auROC of 0.708 and auPR of 0.169. Our model outperformed the baseline model at every age point assessed.

**Figure 3:**
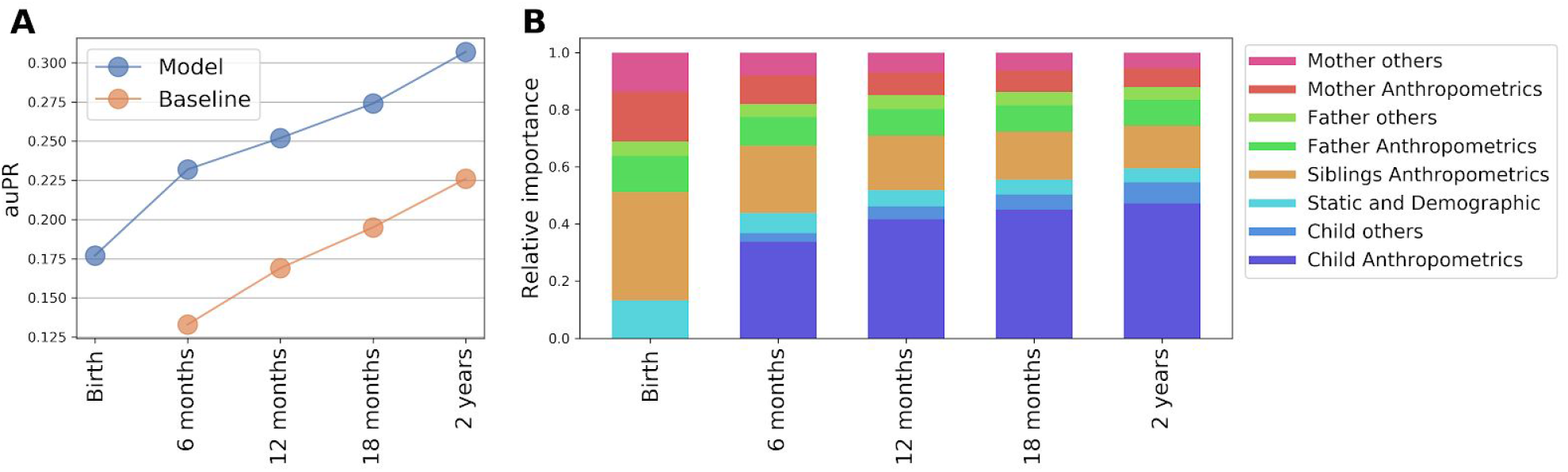
Applying childhood obesity prediction models prior to 2 years of age. **A:** auPR curve for prediction models of obesity at 5-6 years of age based on features that were extracted up to a predefined endpoint age, ranging from birth to 2 years of age. Blue: full model, Red: Baseline model, defined as last routine checkup WFL z-score. **B:** Relative Importance of groups of features for the prediction models, calculated by normalizing to the sum of mean absolute SHAP values for each model. “Others” sums up non-anthropometric or demographic features such as laboratory tests and drugs. Abbreviations: auPR - Area under the PR curve, PR - Precision-Recall, WFL -weight for length

### Feature attribution

An analysis of feature attribution was performed using Shapley values (see Methods). Figure 3A presents a global analysis of the model’s feature attributions. The mean of absolute summation of Shapley values for different groups of features is presented for the entire cohort. We also examined feature importance dependence plots of the Shapley value as a function of the feature value for all individuals (Section 8 in the Supplementary Appendix). As expected, most of the influential features were previous anthropometric measurements of the child, with the last measured WFL percentile being the most impactful feature. We next evaluated the dependence plots for several clinical parameters that have been previously associated with childhood obesity: Familial BMI^31^, ethnicity^32^ and maternal pregnancy glucose levels^33^. Anthropometric features of the child, his parents and siblings **(Fig. 4B, 4D, 4E)** and North African Jewish descendancy **(Fig. 4H)** had a significant impact on the prediction. Maternal blood glucose measured in pregnancy as part of a 50g glucose challenge test (GCT) was also influential for the prediction of obesity, with relative risk increasing monotonically across the entire maternal glucose spectrum and reaching above 1 in values greater than 120 mg/dL **(Fig. 4F)**.

**Figure 4:**
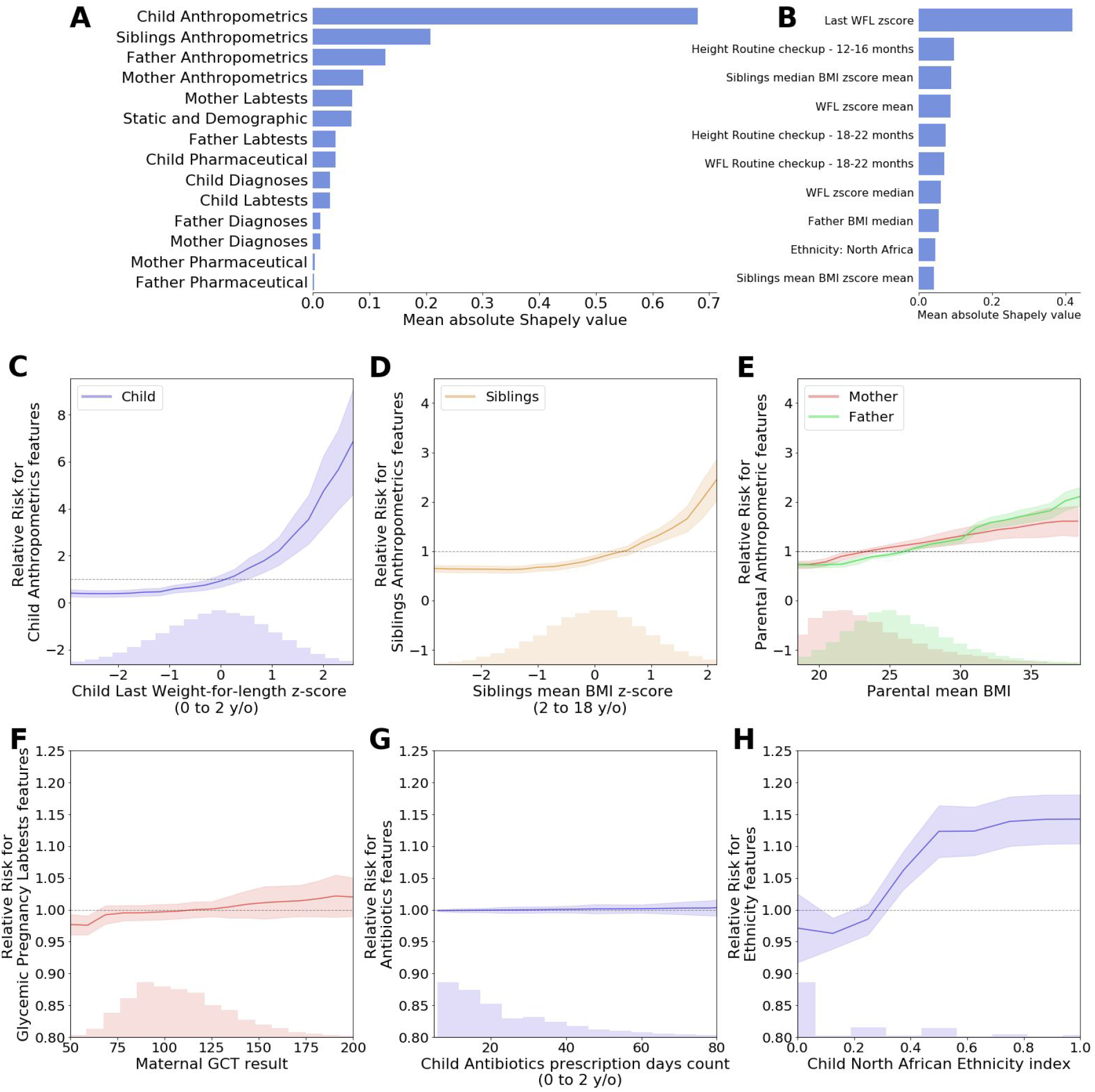
Interpretation of the model. **A**: Shapley values (in absolute log-odds scale) of different groups of features. Note that the scale of the y-axis varies between panels. **B:** Feature importance (Shapley values in log-odds scale) of the top 15 contributing features **C-H**: Plots showing in the lower part a histogram of the distribution of a feature in the data and in the upper part a dependence plot of the predicted relative risk for obesity at 5-6 years of age versus the value of the feature for **C:** child last WFL z-score; **D:** Siblings mean BMI z-score, **E:** Parental mean BMI: Maternal (pink) and Paternal (green); **F**: Maternal 50g GCT results during pregnancy,, **H**: Child North African Ancestry index Abbreviations: GCT- glucose challenge test, WFL- Weight-for-Length, y/o- years old

An analysis of the relative importance of different groups of features at different ages revealed that the most influential features at birth are anthropometric measurements of the siblings, mother and father. Following these, the influence of the child’s own anthropometrics measurements becomes more substantial and is roughly equal to the contribution of all other features at 1 year of age. Laboratory tests, medication dispensing and medical diagnoses have little relative influence, which decreases as the data on the child’s anthropometrics accumulates (**Fig. 3B**).

Using information on pharmaceutical prescriptions, we next analyzed the effect of in utero and early life antibiotic exposure. 28,815 children (22%) were exposed to antibiotics in utero. In pregnancies in which antibiotics were prescribed, there was an average of 1.6 prescriptions and 10.7 prescriptions days per pregnancy. 109,549 children (80%) had at least one antibiotic prescription in the first 2 years of life, with an average of 4.3 prescriptions and 26.5 prescriptions days per child for the children who had at least one prescription. The average age at first antibiotics exposure was 312 days old. Our analysis revealed that the duration of antibiotic exposure in utero and in the first two years of life and the age of first exposure to antibiotics did not impact the prediction of obesity risk at 5-6 years of age **(Fig. 3G)**.

## Discussion

In this study, we developed a prediction model for pediatric obesity at 5-6 years of age based on a comprehensive nationwide EHR encompassing over 10 years of children and familial data. New prevention strategies are crucial to mitigate the rapidly rising prevalence of childhood obesity, especially in light of the limited treatment options available for children who have already developed obesity^35^. Overweight 5-year-olds are four times more likely to become obese later in life compared to normal-weight children^36^, and weight at this age is considered to be a good indicator of the child’s future metabolic health^37^. The target age of our prediction model is also supported by a recently published study on children BMI trajectories^38^. This study, conducted on the German population revealed very similar BMI acceleration patterns as in our population, marking 2 to 6 years of age as the maximal BMI acceleration time period. Our model is therefore timed to identify children at risk prior to this critical time window, in which mature eating patterns become more developed as children reduce breast milk or formula consumption. In addition, our analysis of the transition in obesity status in the first 6 years of life revealed that most obese children had normal weight at infancy, underscoring the importance of building a tool that allows clinicians to identify high risk infants that are considered to have a normal weight at infancy but will develop obesity, as they will constitute the majority of obese children in the future.

Our model achieved an auROC of 0.804 and auPR of 0.307. Further analysis of prediction performance on subpopulations of the cohort demonstrated robustness in discrimination performance across the entire pediatric population, including children with complex chronic diseases. Unlike previous studies^17^, our results were similar for boys and girls. The model aimed at predicting obesity prior to 2 years of age and demonstrated family anthropometric measurements as top features in determining future obesity risk of the child. We have shown that a prediction model constructed pre-birth, which is mainly based on family anthropometric measurements has very similar performance of predicting at 1 year of age based on the child’s last available weight and length measurements **(Fig. 3A)**.

Early identification of children prone to obesity may enable clinicians to implement early lifestyle modifications aimed at obesity prevention. There are several known and potentially modifiable risk factors for childhood obesity including: high energy intake diet, high consumption of sweetened drinks, reduced levels of physical activity and increased time spent in sedentary activities such as television viewing ^39^. Preventive lifestyle modifications can therefore be targeted toward these known risk factors. Although the effect of prevention interventions in early childhood vary across trials, partially resulting from a wide range of interventions tested, some studies show promising results ^14,40,41^. Further prospective and longer term follow up studies are needed in order to identify the optimal prevention strategies for this age group.

Our study has several strengths. First, we employed a sample size of over 130,000 children, which is larger than previous studies on childhood obesity prediction that were based on datasets ranging from 166^42^ to 32,000 children^43^. Second, to the best of our knowledge, our model is the first to include full health care data on both the child, from pregnancy to 5-6 years of age, and the family, and is the first to be validated both temporally and geographically at different clinics on a national level, and represents successful performance on a wide target population. Third, this model is the first to assess clinical utility by constructing decision curves. To date, there are no clinical guidelines defining the risk threshold for obesity prediction. The definition of this threshold may be influenced by many factors, including the characteristics of the proposed intervention, resources available for intervention and the prevalence of obesity in the target population. The threshold will impact the sensitivity and specificity of the prediction model. Decision curve analysis facilitates determining risk thresholds to aid in presenting the performance tradeoffs and thus the clinical relevance of the model for different interventions.

The mechanisms involved in the development of obesity in children are complex and include genetic, developmental, and environmental factors. Our large cohort of Israeli children represents a diverse and multi-ethnic population with genetic heterogeneity^44^. Not surprisingly, many of the variables found to be important in our model were directly related to the child’s previous anthropometric measurements. Familial anthropometric measurements, including paternal, maternal and sibling’s BMI were also important, in line with previous studies showing associations between these variables and childhood obesity^31^. Among familial data, sibling’s BMI had the highest impact on the prediction model, most likely due to shared genetic and environmental influences.

There is evidence that the uterine environment may cause a permanent influence on fetus’ future health, and may lead to enhanced susceptibility to diseases later in life. This concept is defined as ‘gestational programming’, and is thought to be mediated by epigenetic mechanisms^45,46^. Data on maternal pregnancy, allow a unique opportunity to analyze associations of these features to the future obesity status of the offspring. Interestingly, one of the top influential features in pregnancy was maternal blood glucose values (**Fig. 4F**). An increase in GCT during pregnancy, adjusted for all other features incorporated in the model (such as maternal BMI), was associated with a higher risk for childhood obesity. This association, which was apparent even in glucose values which are considered in the normal range, is in line with recent studies, demonstrating that exposure to higher glucose levels in utero throughout the entire maternal glucose spectrum is significantly associated with childhood glucose and insulin resistance of the offspring and is independently associated with childhood adiposity^33,47,48,49^. Ethnicity as a risk factor has previously been studied in the UK and USA populations, in which a higher prevalence of obesity was found among children of African descent^32^. In the Israeli population, our analysis revealed North African Jewish descendancy as a contributor for predicting obesity.

Our study also has several limitations. First, the dataset does not contain information on nutrition or lifestyle habits, parental education and socioeconomic status. Data on mode of delivery, gestational age and birthweight were available for only a portion of the children included in the cohort. In addition, we did not have genetic information in our possession. Although a previous study demonstrated that currently known genetic variants have a relatively small contribution in childhood obesity prediction^54^, a recent genome-wide polygenic risk score for obesity was associated with a gradient in weight that started to emerge in early childhood, and predicted differences in weight during early childhood^55^ .Thus, the incorporation of genetic information to the model may further enhance its predictive performance. Second, our data is retrospective in nature and thus may suffer from potential biases and be affected by a variety of healthcare processes^56^. We tried to minimize sampling bias by choosing children based on the schedule of routine measurements of weight and height. Third, the prediction model is based solely on data of Israeli children. However, our validation process, including a geographical validation, the well-known universal risk factors for childhood obesity emerging from our analysis of the model, and the striking similarity of our analysis on BMI trajectories to an independent, recently published German cohort^38^, all indicate that our results may be generalized to other populations as well. Finally, although the first study to explore the use of obesity risk tools in clinical practice demonstrated that identification of future risk during infancy was viewed positively by parents^57^, no studies have examined the clinical impact of using these tools for intervention. Our study paves the way for future trials of focused intervention addressing the real life efficacy of this approach.

Overall, our large-scale national risk assessment model is an important step in prioritizing prevention strategies for childhood obesity. Childhood obesity can be accurately predicted during infancy or even at birth.The reported model may have a direct and immediate impact on large populations of children, and can be embedded into current health services systems without additional costs.

## Data Availability

The data that support the findings of this study originates from Clalit healthcare. Restrictions apply to the availability of these data and so are not publicly available. Due to restrictions, it can only be accessed through requests from the authors and/or the Clalit healthcare organization.

## Supplementary Appendix

### 1. Clalit EHR

Clalit is the largest of the four health care organizations in Israel and one of the largest in the world, serving over 4.4 million individuals (over half of the Israeli population). Israel’s adoption of Electronic Health Records (EHR) started during the mid-1990’s and provision of healthcare services in both the primary care and hospital setting have been recorded in EHRs by all care professionals in hospitals and community for more than 20 years. Clalit has 100% (single software) EHR coverage and data all feeds into an aggregated data warehouse. The Clalit member population is stable from birth to death (< 2% annual turnover) and the data is longitudinal (spans from birth to death) and includes both claims and direct clinical data from nearly all clinical and health care interactions. The data are linked through a unique identifier and geo-coded with the patient’s clinic and home address, and aligned with area level statistics. The data warehouse uses a single, universally adopted EHR system throughout the entire organization. Data are reflective of the members’ full clinical experience, capturing administrative, financial and clinical information across hospitals (inpatient and emergency department settings), primary care clinics, specialty clinics, pharmacies, laboratories, clinical measures, diagnostic and imaging centers, allied health services, dental, and complementary health services, as well as organizational and national disease registries. The data are unique not only in their breadth and historical depth, but also that they are harmonized (single EMR software), and considered highly accurate, as the lack of coding-based payment system (i.e. DRG) in Israel generates upcoding-free clinical data. The data warehouse is also linked to national databases providing socio-demographic information related to patients and clinics. Clalit owns and operates approximately 1,500 primary care clinics and 14 hospitals, including 30% of Israel’s hospital acute care beds.

### 2. Cohort selection

**Figure S1:**
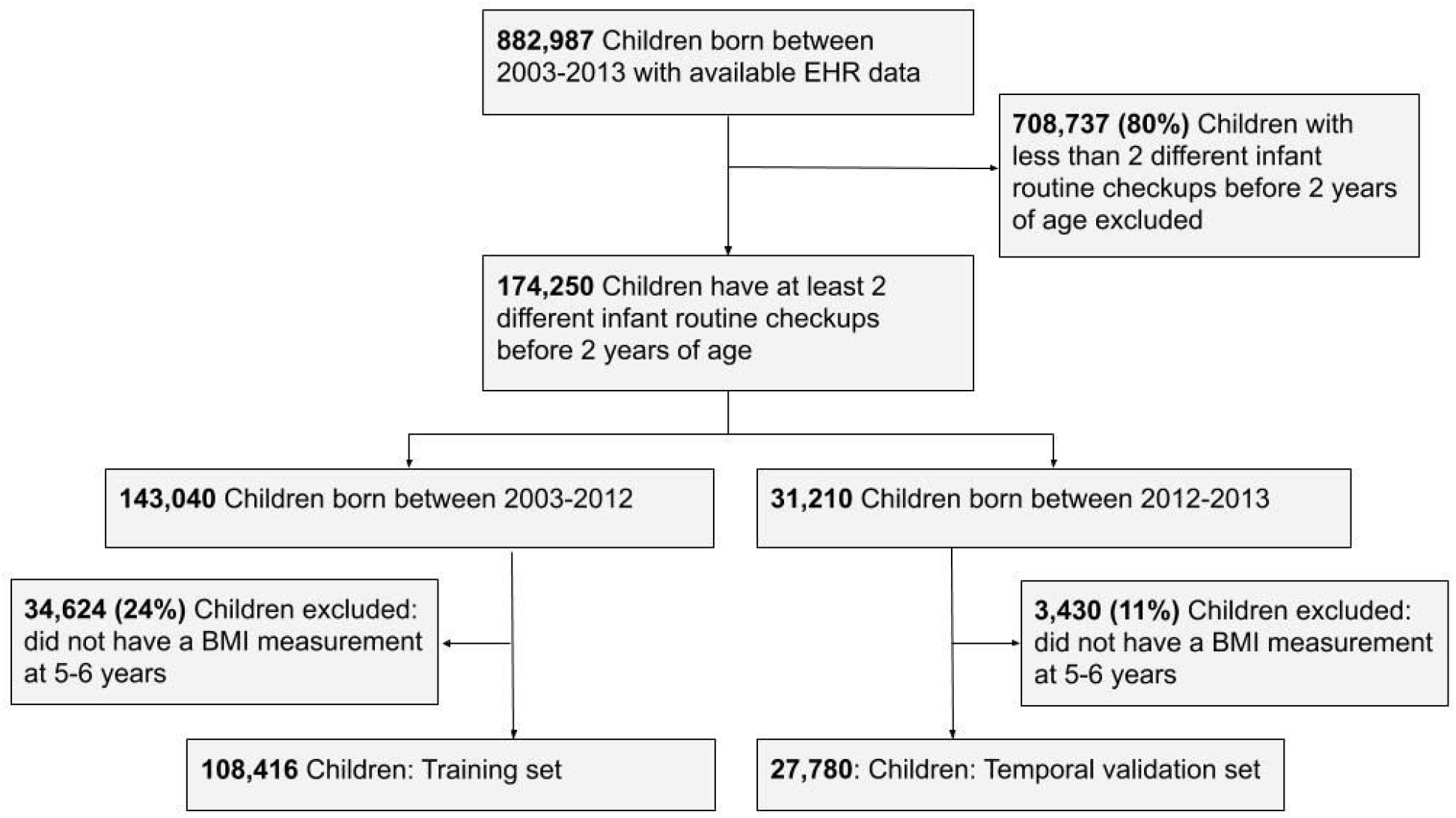
Cohort selection. Abbreviations: EHR- Electronic health record

### 3. Features generation methods

A total of 1,556 features were constructed from the dataset. The following list describes the generation mechanism for each. We defined “index date” as the date of applying prediction. All features were taken prior to the index date.

A. Child’s features:
  a. Demographics :
    i. Sex
    ii. Born in Israel (True/False)
    iii. Date of birth
    iv. Week of year (1-52) at birth
    v. Features describing ancestry: 15 features breaking down the origin of the patient’s ancestors, as logged in their country of origin. World’s countries were clustered into 14 categories, corresponding to Israel’s major ancestry groups: North Africa, Iraq, Iran, Yemen, East Europe, West Europe, ex-USSR, North America, Latin America, Arab, Mediterranean, Ethiopia, Asia and Africa. Another feature logs the percentage of unknown origin. The distribution of the different ancestries indices in the cohort is presented in Fig. S2.

**Figure S2:**
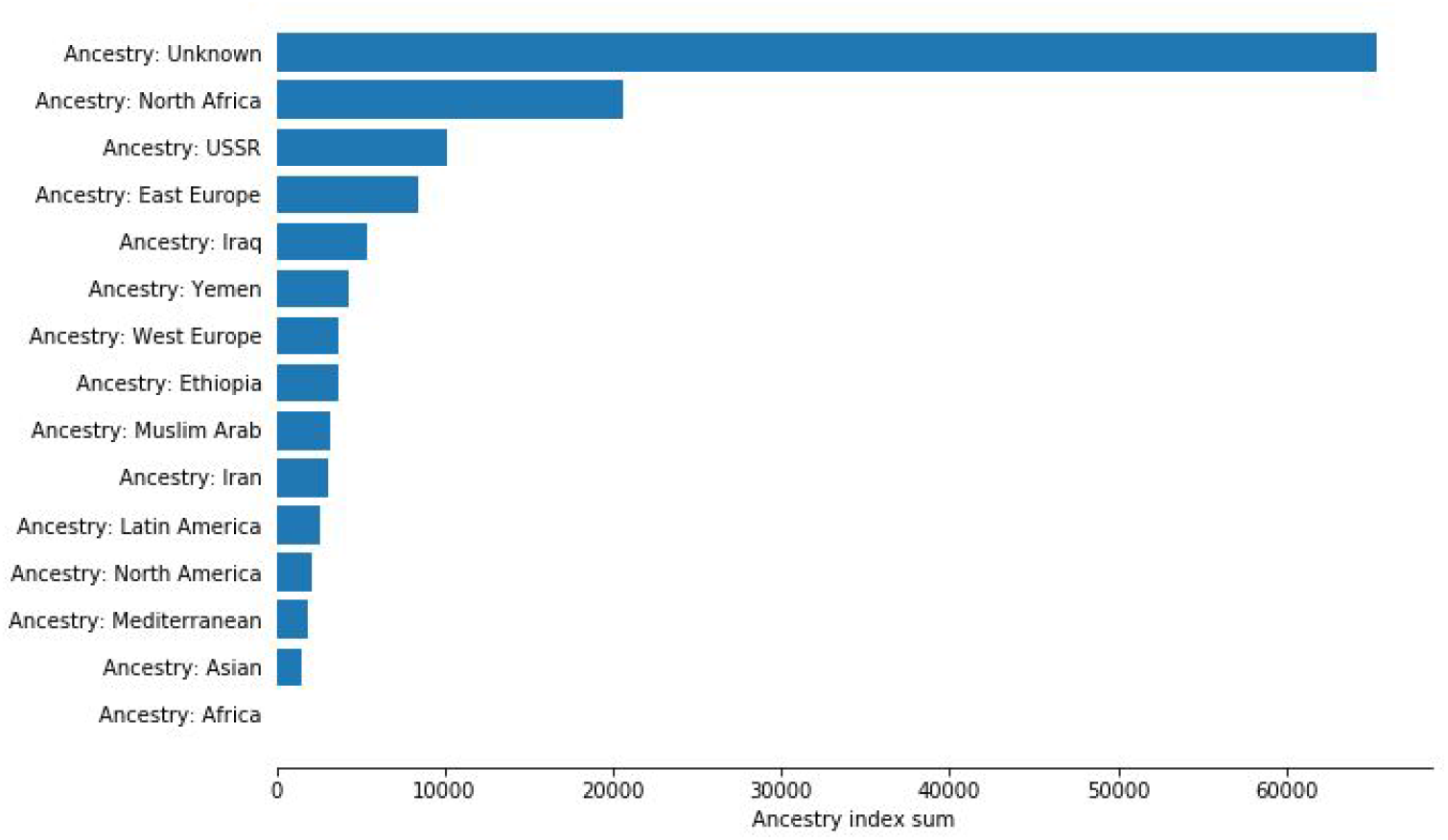
The distribution of the different ancestries index in the cohort.
    vi. Socio-economic data: Although personal socio-economic data were not available, we generated estimates using the locality of most of the patient’s clinic visits and data available by Israel’s Central Bureau of Statistics. Features include locality type (length 20, 1-hot vector) and locality religion breakdown (length 5, summing to 1 vector).
    vii. Number of known siblings
    viii. For children born as part of a multiple gestation, number of offspring born
    ix. Birth order
B. Birth features (data available for children born in a Clalit hospital after 2012):
  i. Birthweight
  ii. Premature birth label from hospital record
  iii. Mode of delivery: Cesarean delivery
  iv. Epidural analgesia at birth
C. Anthropometrics measurements: Weight and height measurements were used for BMI and Weight-for-Length (WFL) percentile calculations. In Israel, Infant routine checkups are done in special centers called “Tipat Halav”. In these centers, routine measurements of weight and length are performed in predefined time windows: 1,2,4,6,9,12 and 18 months. These centers are distributed throughout Israel and are operated by the health bureaus. The service at “Tipat Halav” centers is provided by various parties: Ministry of Health, the health maintenance organization and the municipalities (depending of the place of residence). In this study, we only had “Tipat Halav” data from the Clalit health organization at our disposal. We chose to analyze anthropometric data gathered during the routine checkups in order to reduce selection bias that may originate from including non-routine measurements. This bias can be seen empirically when plotting our weight and height measurements data against the CDC percentile curves, as an average lower BMI in times in which routine checkups are being held(**Fig. S3**).

**Figure S3:**
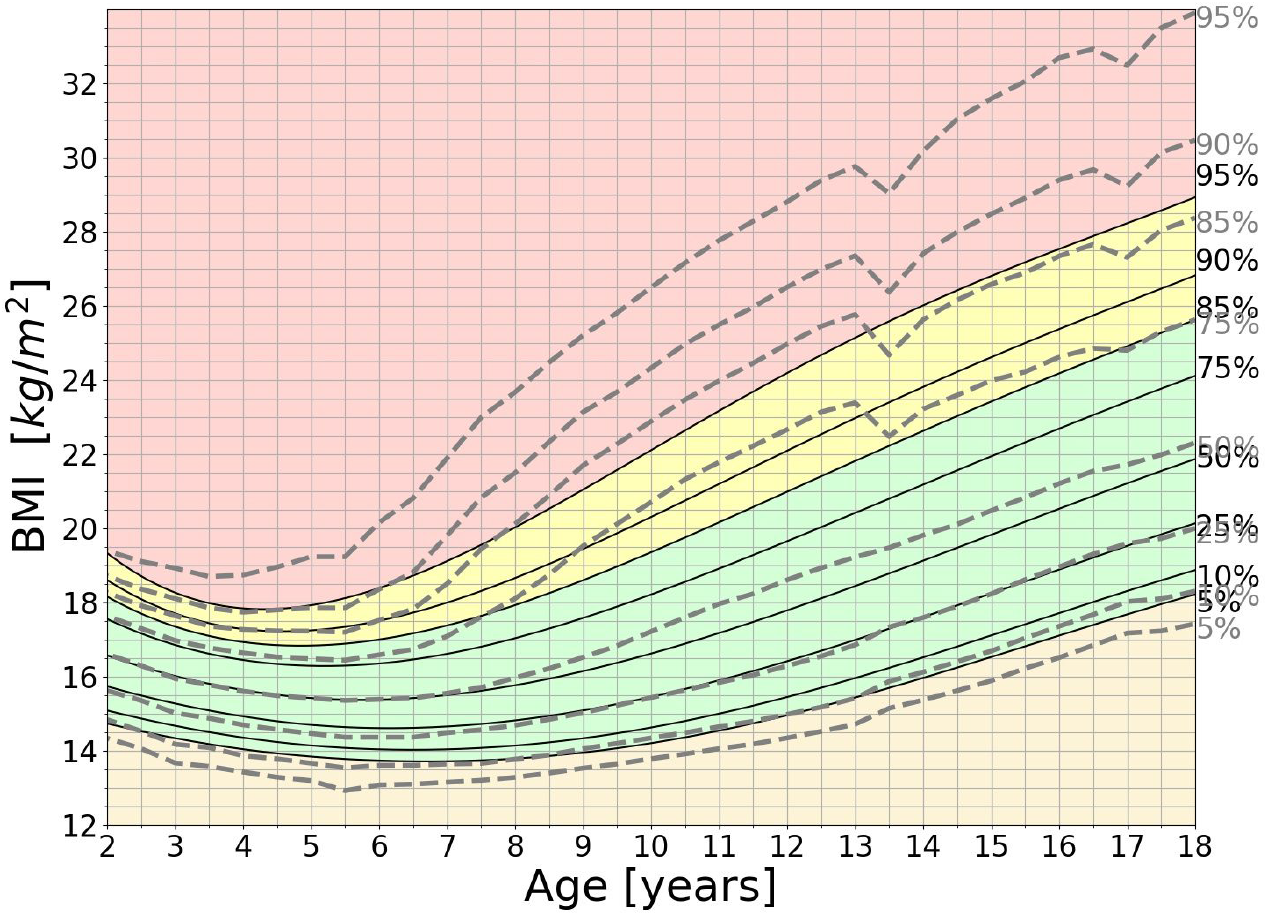
BMI percentiles by child age. Solid lines are CDC growth charts. Dashed lines are calculated percentiles from our database. “Less biased” measurements can be seen at the routine checkups made at ages 5, 13, 17. Abbreviations: BMI - body mass index, CDC- Center for Disease Control and Prevention We therefore first binned the weight and length data with accordance to the routine checkups schedule. We defined a *base-rate* of expected measurements per age at primary care setting (horizontal red line in **Fig. S4 B**). The time range around each infant routine checkup was estimated by periods in which the empirical measurements rate was higher than the base rate. Measurements during these time ranges were considered as routine measurements. An analysis of missing anthropometric data in the routine checkups is presented in **Fig. S5**. To further mitigate the probability of selecting children who did not come at a routine checkup, we only included children having at least 2 estimated routine measurements in separate bins in our study.

**Figure S4:**
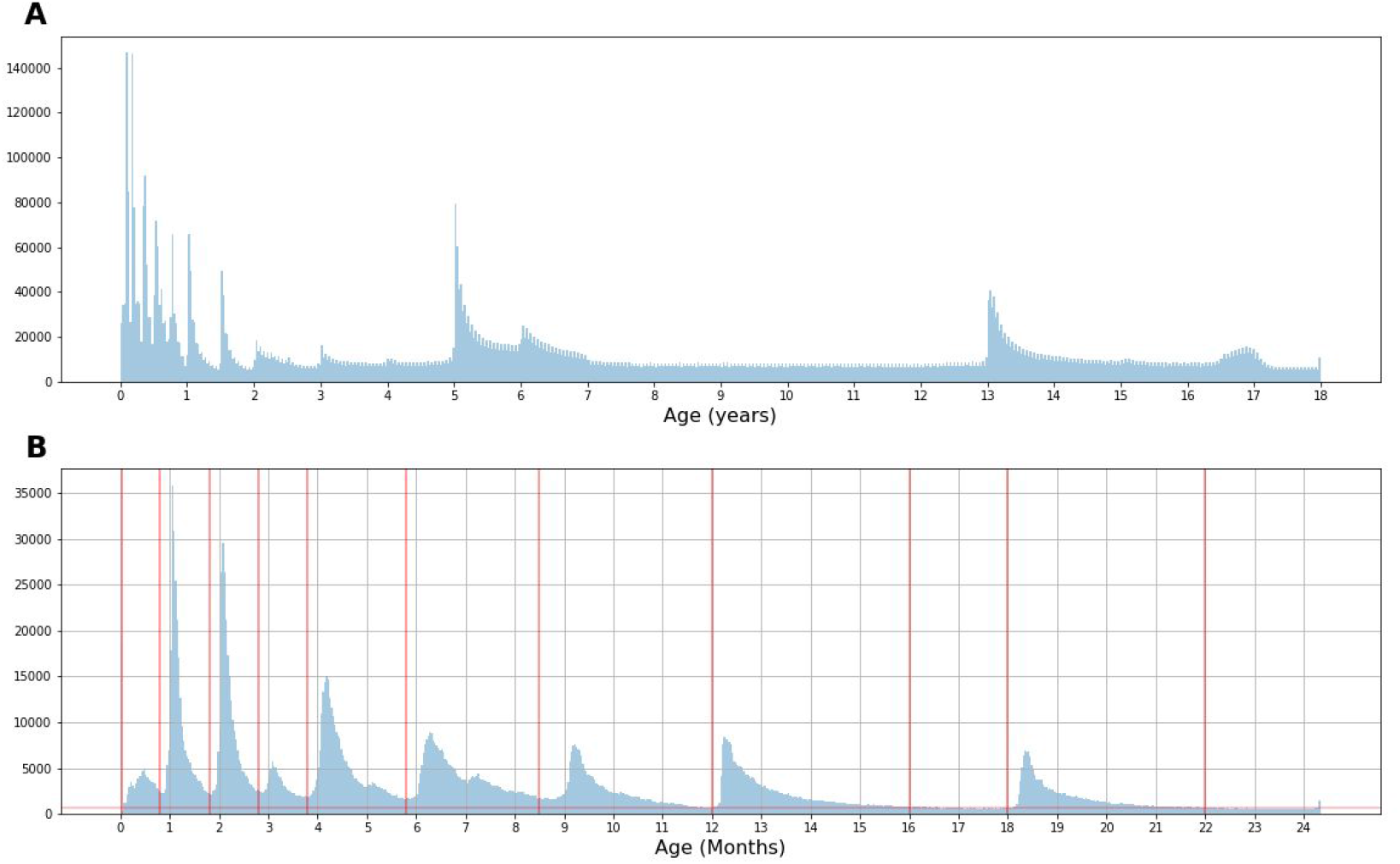
Empirical histograms for weight and height measurements. **A:** Histogram of all BMI measurements up to 18 years of age. **B:** Infant routine checkup windows for our cohort. Abbreviations: BMI - body mass index

**Figure S5:**
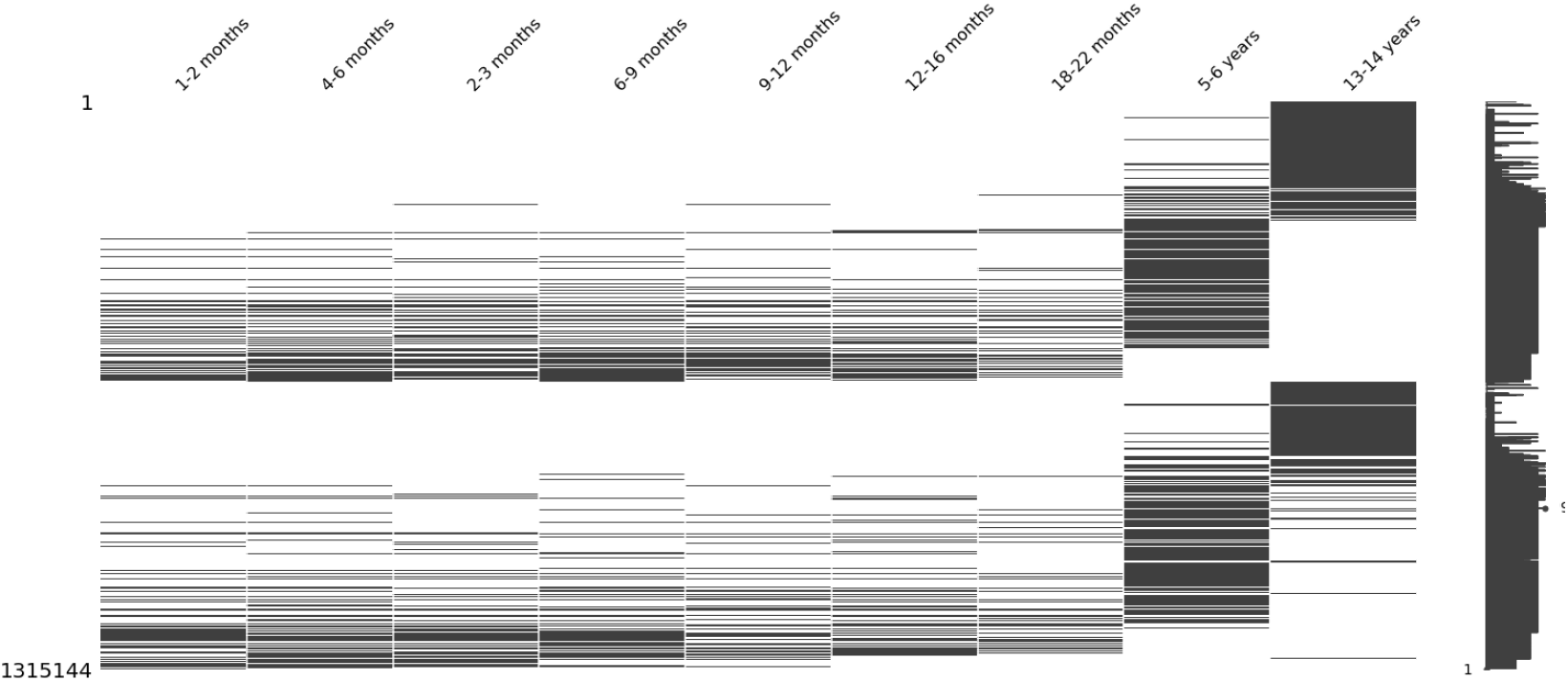
**Existing\missing data pattern** for weight and height routine checkups at 1,2,4,6,9,12, 18 months and 5,13 years of age. For each infant routine checkup bin, the weight, height and derived WFL or BMI measurements were converted to CDC z-scores (adjusted for age and sex) and these were taken as features for the model. Apart from the routine checkups, all WFL and BMI z-score measurements prior to the index date were summarized by the following statistics: mean, median, maximum, minimum, standard deviation and count. Slope and intercept of a linear fit for all WFL z-score measurements, and last available WFL z-score and age at that last measurement were also used as features.
D. Laboratory tests: Mean values of:
  1. Routine blood tests. In Israel, there is a routine blood test for Complete Blood Count (CBC) for children at the age of 9-12 months in order to screen for Iron**-**deficiency Anemia.
  2. Blood tests which were available for at least 1% of children (total of 50 blood tests).
E. Clinic and hospital diagnoses: Diagnoses in the form of ICD9 codes were manually sorted by a pediatrician. Overall, 148 Diagnoses were included in the analyses. Diagnoses that were available for less than 1% of children in the cohort and diagnoses with no medical implications such as “Observation” or “Fill out forms” were excluded from the analysis.
F. Medications: Drugs purchased were classified according to ATC5 categories and were manually sorted by a pediatrician. Overall, 28 ATC5 categories were included in the analyses. Drugs that were purchased for less than 1% of the children in the cohort, over-the-counter drugs and drugs administered locally (such as skin ointments and ophthalmic eye drops) were not included as features. Systemic antibiotics purchased were classified to one of the following categories: Penicillin, Beta lactam Penicillin, Macrolides, Cephalosporin and Sulfonamides. Features of antibiotic usage included a count of the total number of days in which the child was prescribed with antibiotics and the age of first antibiotics prescription.
G. Maternal Features:
  1. Anthropometrics measurements and vital signs were divided into 4 bins: Pre-pregnancy (defined as the two years prior to pregnancy initiation) and 3 trimesters of pregnancy; first trimester was defined as up to 14 weeks of gestation, second trimester was defined as 14-28 weeks of gestation and third trimester was defined as week 29 of gestation to delivery. Trimesters dates were estimated as following: For children born in a Clalit hospital after 2012, gestational age was available and was used for approximation; For women with available GCT results, performed routinely in Israel at 24-28 weeks of gestation, GCT result was used as a proxy for week 26 of gestation and then used for approximation; Otherwise, the date of delivery was considered as week 40 of gestation and then used for approximation.
    i. Weight and height measurements were used for calculation of mean BMI and BMI differences between time periods. Other statistical measures, including mean, median, minimum, maximum, standard deviation and count from the Pre-pregnancy period were also used as features.
    ii. Systolic and Diastolic Blood pressure mean values.
  2. Laboratory tests were also divided into 4 bins: Pre-pregnancy and 3 trimesters of pregnancy as described above. Mean values in each bin were used as features Blood tests which were available for at least 1% of mothers (total of 439 features).
  3. Clinic and hospital diagnoses: Diagnoses in the form of ICD9 codes during pregnancy were manually sorted by a physician. Overall, 112 diagnoses pre-pregnancy and 76 diagnoses during pregnancy were included in the analyses. Diagnoses that were available for less than 1% of the women in the cohort and diagnoses that had no medical implications were excluded from the analysis.
  4. Medications: Drugs purchased were classified according to ATC5 categories and were manually sorted by a physician. Overall, 65 ATC5 categories were included in the analyses. Drugs that were purchased by less than 1% of the women in the cohort, over-the-counter drugs and drugs administered locally were not included as features.
H. Paternal features were extracted from 2 years prior to the delivery of the child to the index date.
  1. Anthropometric measurements: Weight, height and BMI measurements were used as features. Statistical measures, including mean, median, minimum, maximum, standard deviation and count were of these were derived and used as features.
  2. Laboratory tests: Mean value (89 laboratory tests that were available for more than 1%).
  3. Clinic and hospital diagnoses: Diagnoses in the form of ICD9 codes were manually sorted by a physician. Overall, 384 Diagnoses were included in the analyses.
  4. Diagnoses that were available for less than 1% of the fathers in the cohort and diagnoses that had no medical implications were excluded from the analysis.
  5. Medications: Drugs purchased were classified according to ATC5 categories and were manually sorted by a physician. Drugs that were purchased by less than 1% of the fathers in the cohort, over-the-counter drugs and drugs administered locally were not included as features. Drugs that may indicate a paternal comorbidity that may be associated with obesity, including anti diabetic drugs, antihypertensive drugs and antilipemic drugs were included. Overall, 17 ATC5 categories were included in the analyses.
I. Siblings features:
  a. Anthropometric measurements: BMI z-score statistics, including mean, median, maximum, minimum, standard deviation, and count over all of the child’s siblings. Mean and standard deviation of BMI z-score at time windows at 5-6 and 13-14 years of age were taken as features as well.

### 4. Distribution of obesity status at the target age

**Table S1.** Obesity status distribution at the target age.

| Table S1. Obesity status distribution at the target age |  |  |  |  |
| --- | --- | --- | --- | --- |
|  |  | Train set (n=108,416)<br>aged 5 years old before<br>2017 | Temporal test set<br>(n=27,780) aged 5<br>years old at 2017 | All (n = 136,196) |
| Obesity status at<br>5-6 years of age | Underweight | 13,635 | 3,304 | 16,939 |
|  | Normal weight | 75,648 | 19,867 | 95,515 |
|  | Overweight | 19,133 | 4,609 | 23,742 |
|  | Obese | 8,120 | 1,941 | 10,061 |

### 5. Prediction model parameters

We used a gradient boosting trees model trained with the LightGBM ^58^ python package. Hyperparameters were selected following a cross-validated grid search, with the following settings selected:

- num_boost_round = 500
- num_leaves = 24
- feature_fraction = 0.4
- bagging_fraction = 0.8
- bagging_freq = 5
- min_data_in_leaf = 80
- min_data_in_bin = 3
- max_bin = 128

Missing values were not imputed prior to training the model. LightGBM ^58^ uses the method of *block propagation* in which tree splits are learned only from non-missing data, and only after that, the direction of splitting missing values is learned (by minimizing error) for the whole block of samples for missing values for that feature. Josse et al has shown that this procedure is a good option relative to various imputation methods ^59^.

### 6. Model discrimination performances on specific subgroups

**Figure S6:**
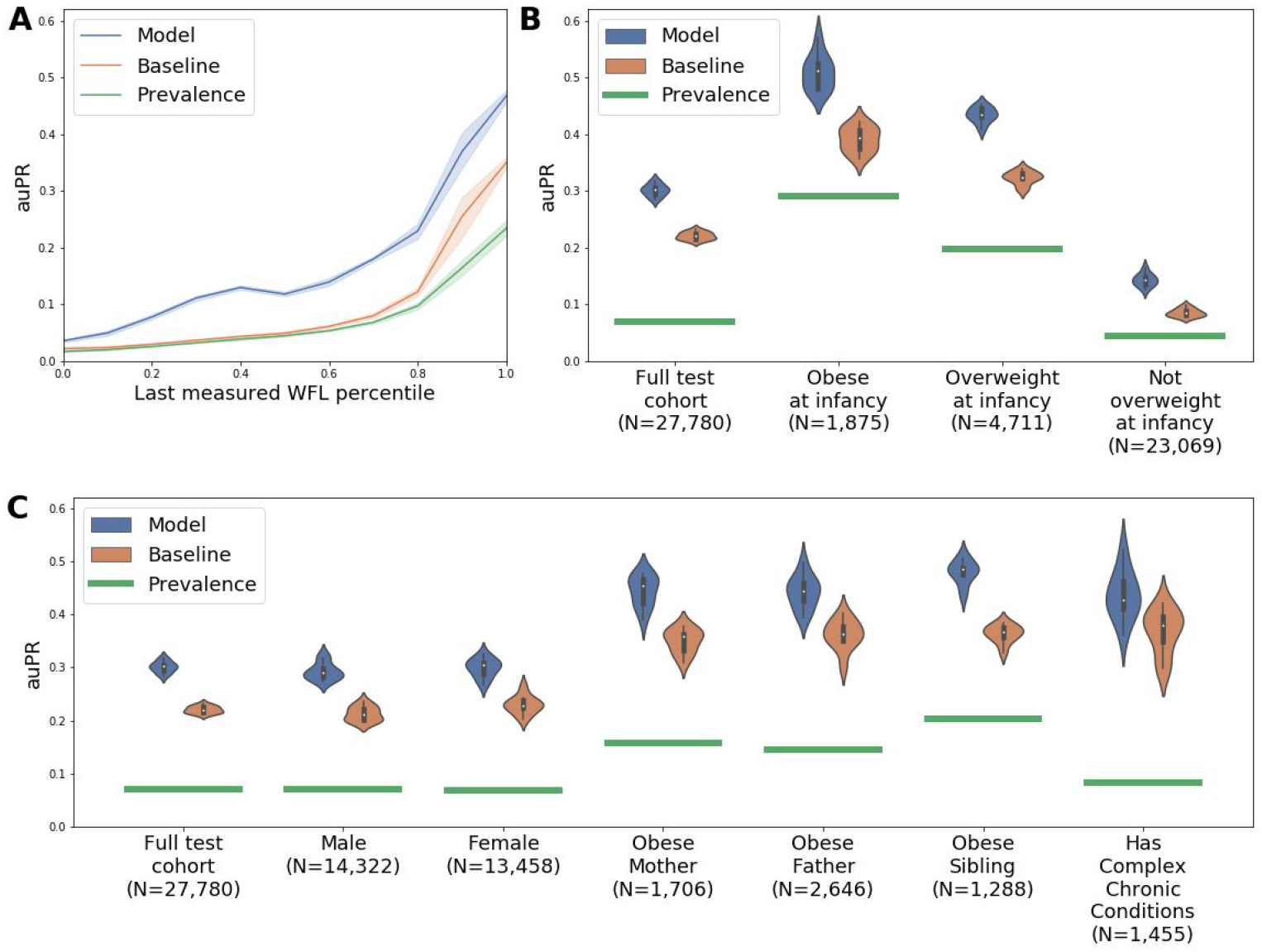
**Model discrimination performances** represented by Precision-Recall (auPR) according to: **A**:Last measured WFL percentile ;**B:** Obesity status defined by the last infant routine checkup; **C:** Different subpopulations of children Abbreviations: auPR - Area under the PR curve, PR - Precision-Recall, WFL - weight for length

Model discrimination performances as a function of the number of available infant routine checkups:

**Figure S7:**
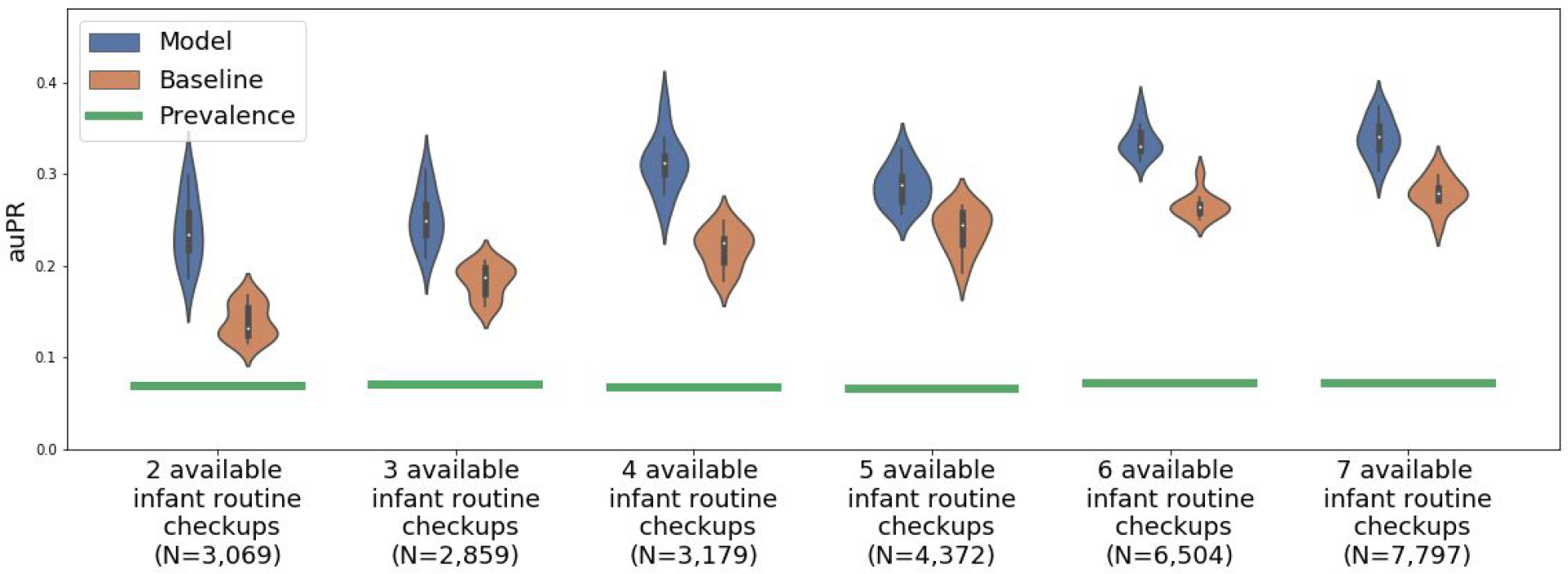
Model discrimination performances represented by Precision-Recall (auPR) according to the number of documented anthropometric measurements during routine checkups.

### 7. Applying childhood obesity prediction models prior to 2 years of age

**Table S2.**
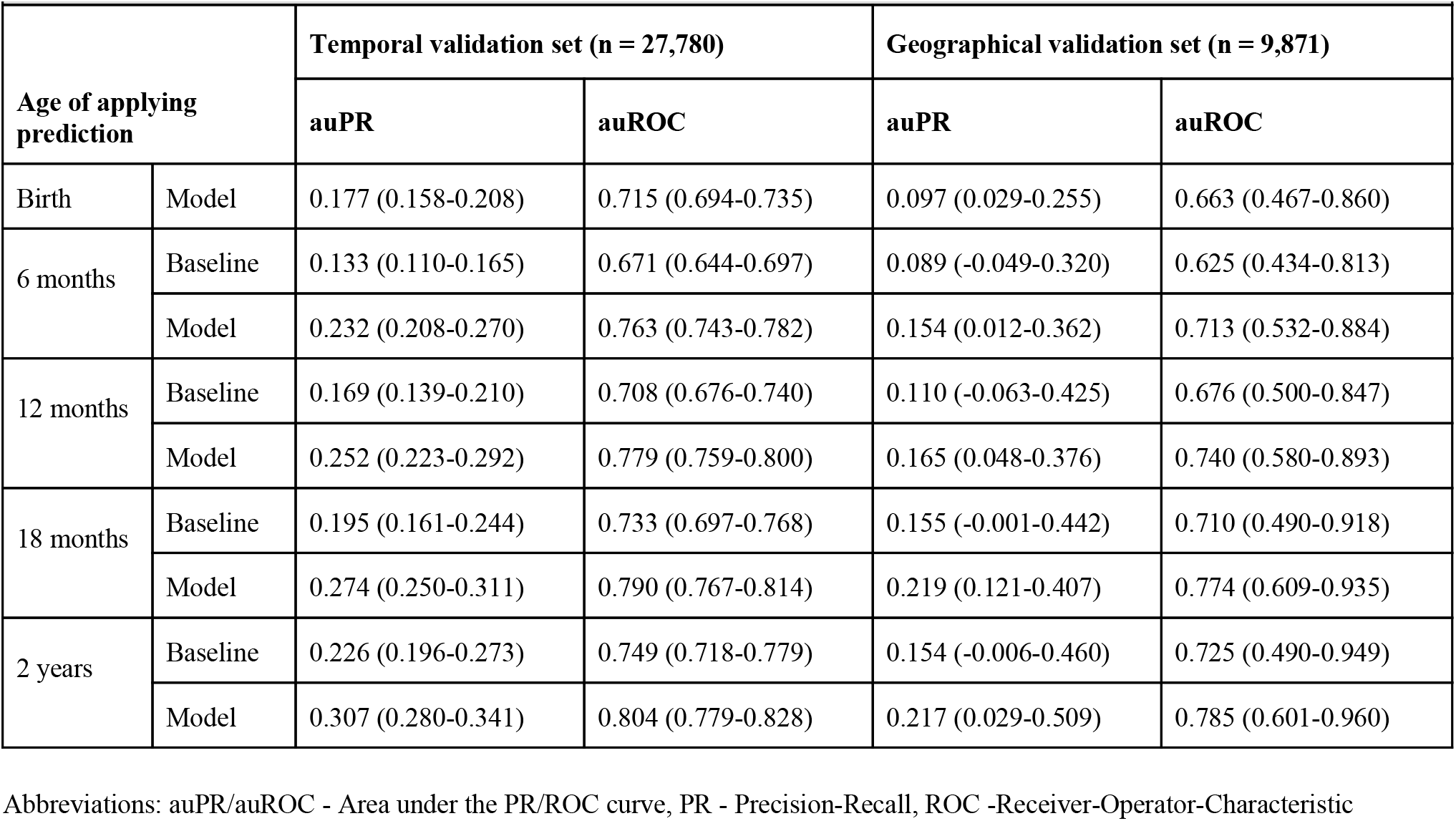
Prediction of obesity at 5-6 years of age depending on age of applying prediction.

### 8. Feature attributions by SHAP

#### Individual feature attributions by SHAP

SHAP values are calculated per feature per patient, which therefore enables an analysis of individual features contribution. A caregiver can use the individual SHAP analysis to gain insights on a specific patient risk prediction and identify which personal clinical parameters were most influential for that prediction. Below is an example of such an analysis for a specific child in the cohort (**Fig. S8**). The model outputs an obesity risk prediction of 83% for this specific child. A caregiver can then evaluate that the most influential features increasing the predicted risk of the child were the child’s own anthropometric measurements while his siblings BMI decreased the predicted risk for obesity.

**Figure S8:**
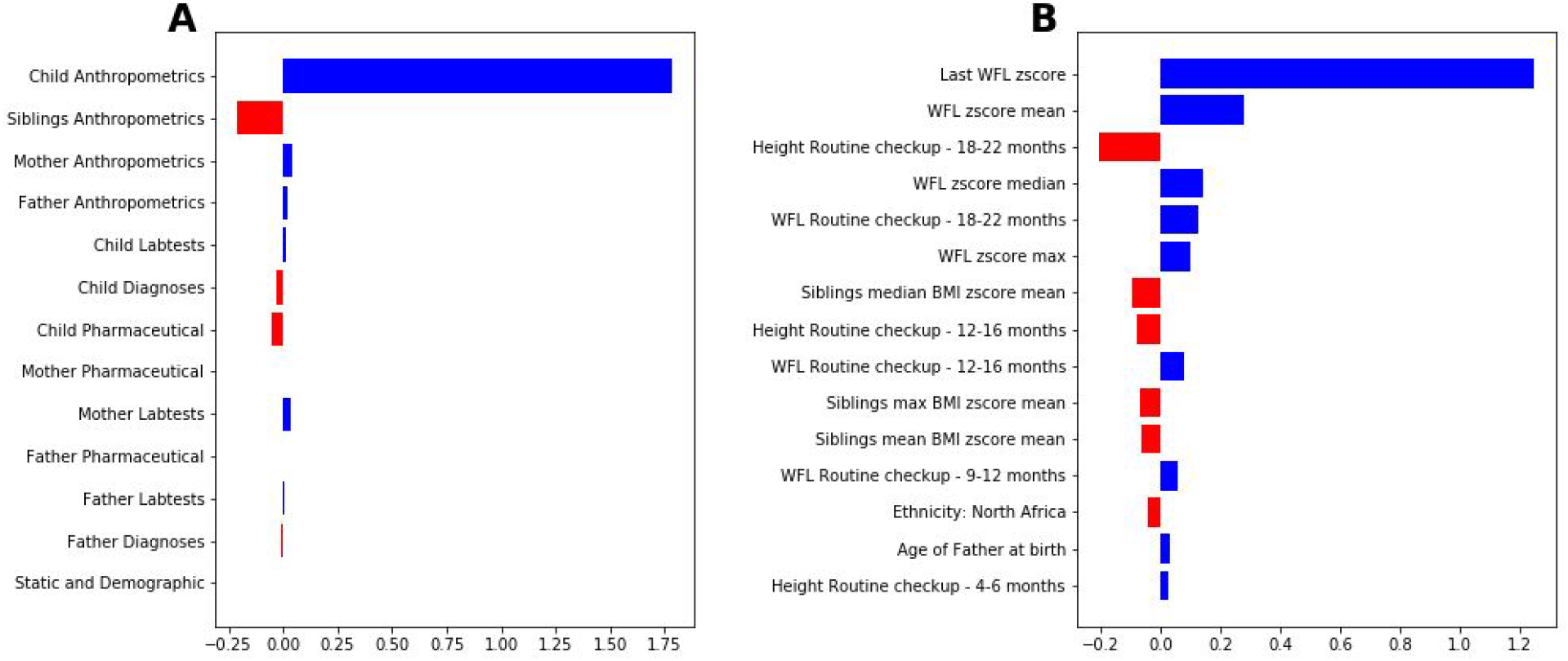
Analysis of Individual feature attributions by SHAP. **A:** Shapley values (in log-odds scale) of the obesity prediction model divided to different groups of features for a specific child in the cohort. The summation of all values equals the individual log-odds of obesity risk **B:** Shapley values (in log-odds scale) of the top 15 contributing features to the specific child prediction. Abbreviations: BMI - body mass index, WFL- Weight-for-Length

### Dependence plots and relative risk score

Shapley values are calculated individually for every child’s feature. An important property of Shapley values is that they are additive, meaning that the Shapley values of a child’s features add up to the predicted log-odds of obesity for that child.

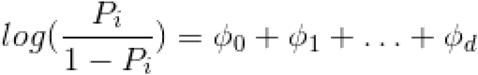

where *ϕ* _0_ is defined as the “bias” Shapley value (the logit of the population prevalence *P*_0_), and *ϕ* _*i*_ for *i* ∈ {1, …, *d*}are the Shapley values related to features 1, …, *d*. The predicted probability based on the contribution of a single feature is then:

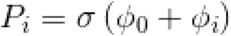

Where *σ*(*x*) is the inverse of the logit function - the Sigmoid function.

We further transform this value for each feature and each child and obtain a *SHAP Relative Risk score* (RR) defined below:

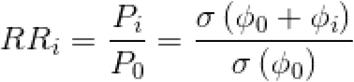

Building a model with many correlated features (e.g. a child’s weight measurement at adjacent timepoints) is bound to suffer from severe collinearity of the features, and consequently the feature attributions will be spread across these related features. To tackle this, we made use of the additive property of Shapley values. By adding up the Shapely values of related features, we can report an analysis on this group of features. This gives better estimates of relevant risk scores per feature category. For a set *D*={*i, j*, …},of features, the *SHAP Relative Risk score* definition extends to

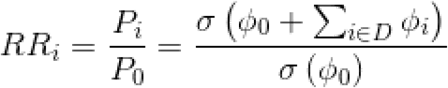

### 9. Clinical utility analysis

Decision curves ^25^ offer a graphical tool to analyze clinical utility of adopting a new risk prediction model. The curves contain information that can guide clinicians to make decisions based on the model’s outputs, and based on the consequences (costs and benefits) of their decision to treat.

We follow the TRIPOD (Transparent Reporting of a multivariable prediction model for Individual Prognosis Or Diagnosis) guidelines ^60^ that recommend calculating and plotting decision curves ^25^ based on the following Net Benefit (*NB*) calculation:

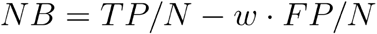

Where *TP, FP* are the number of true positives and false positives for a given threshold probability respectively, *N* is the total number of samples and *w* is the proportion between the cost, *C* of a false positive and the benefit, of a true positive. If we demand a zero or positive net benefit, *B* it can be shown [29] that *w* is connected to an optimal threshold probability by:

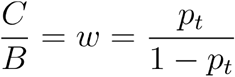

As a simulated real-world example, assume a hypothetical situation in which the health service provider decides to provide nutritional and lifestyle guidance programs to families of children at risk of being overweight in the future. Due to clinical and economical reasons, the benefit and cost of this treatment are expressed in the following way : For every 10 children that will undergo the guidance program, the health service is willing to squander the costs of “mistakenly” guiding 9 children who will not be overweight in the future, provided that one of the ten is a true future overweight child who may actually benefit from this program. This description can now be translated into an optimal probability threshold of *p*_*t*_ = 0.1.. The health service provider can now refer to the decision curve presented in **Fig. S9** and observe that at this value the net benefit of using our model over other treatment strategies is higher, and also quantify the net benefit difference between using our model and other models and strategies such as: treat none, treat everyone and treat by baseline model.

**Figure S9:**
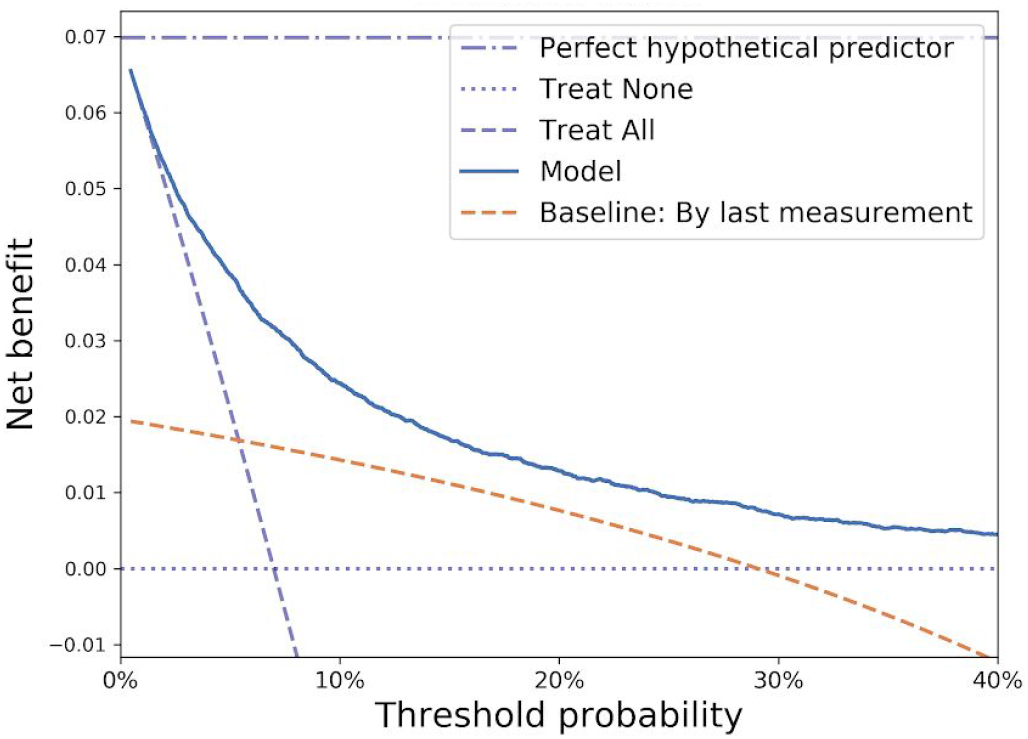
**Decision curve analysis** containing different treatment strategies of our model (blue) and the Baseline model (red). Strategies of treating all, treating none and the perfect hypothetical predictor are also presented.

## Data Availability

The data that support the findings of this study originates from Clalit healthcare. Restrictions apply to the availability of these data and so are not publicly available.

## Code availability

The code that supports the findings of this study is tailored to the data and the fields of the Clalit healthcare database and is thus not provided since it is of no use as a standalone without access to the data itself. The algorithmic models used Python code packages which are publicly available.

## Acknowledgements

We thank Gabi Barabash, Elad Barkan, Iris Kalka and members of the Segal group for discussions. E.S. is supported by the Crown Human Genome Center; D. L. Schwarz; J. N. Halpern; L. Steinberg; and grants funded by the European Research Council and the Israel Science Foundation

## Author information

These authors contributed equally: H. Rossman, S. Shilo, S. Barbash-Hazan

### Affiliations

*Department of Computer Science and Applied Mathematics and Department of Molecular Cell Biology, Weizmann Institute of Science, Rehovot, Israel*

Hagai Rossman, Smadar Shilo, Nitzan Shalom Artzi, Eran Segal

*Pediatric Diabetes unit, Ruth Rappaport Children’s Hospital of Haifa, Rambam Healthcare Campus, Haifa, Israel*

Smadar Shilo

*Helen Schneider Hospital for Women, Rabin Medical Center, Petach Tikva, Israel*

Shiri Barbash-Hazan, Eran Hadar, Arnon Wiznitzer

*Sackler Faculty of Medicine, Tel Aviv University, Tel Aviv, Israel*

Shiri Barbash-Hazan, Eran Hadar, Arnon Wiznitzer

*Clalit Health Services, Clalit Research Institute, Tel Aviv, Israel*

Ran D. Balicer, Becca Feldman

*Department of Epidemiology, Faculty of Health Sciences, Ben Gurion University, Beer sheva, Israel*

Ran D. Balicer

## Contributions

H.R., S.S. and S.B.H conceived the project, designed and conducted the analyses, interpreted the results and wrote the manuscript and are listed in random order. N.A. conducted the analyses and wrote the manuscript, E.H., R.D.B and B.F. interpreted the results. A.W. and E.S. conceived and directed the project and analyses, designed the analyses, interpreted the results, wrote the manuscript, and supervised the project.

## Corresponding authors

Correspondence to Arnon Wiznitzer and Eran Segal

## Ethics declarations

